# Binary toxin expression by *Clostridioides difficile* is associated with worse disease

**DOI:** 10.1101/2021.07.30.21261256

**Authors:** Mary K. Young, Jhansi L. Leslie, Gregory R. Madden, David M. Lyerly, Robert J. Carman, David B. Stewart, Mayuresh M. Abhyankar, William A. Petri

## Abstract

**Background:** The incidence of Clostridioides difficile infection (CDI) has increased over the past two decades and is considered an urgent threat by the Centers for Disease Control. Hypervirulent strains such as ribotype 027, that possess genes for an additional toxin *C. difficile* binary toxin (CDT) are contributing to increased morbidity and mortality. In the mouse model, CDT activates Toll-like receptor 2 resulting in suppression of a protective type 2 innate immune response mediated by eosinophils.

**Methods:** We retrospectively tested stool from 215 CDI patients for CDT by enzyme-linked immunosorbent assay (ELISA). Stratifying patients by CDT status, we assessed if disease severity and clinical outcomes correlated with CDT positivity. Additionally, we performed 16 S rRNA gene sequencing to examine if CDT positive samples had an altered fecal microbiota.

**Results:** We found that patients with CdtB, the pore forming component of CDT, detected in their stool were more likely to have severe disease and had higher 90-day mortality compared to CDT negative patients. CDT positive patients also had higher *C. difficile* bacterial burden and white blood cell counts. There was no significant difference in gut microbiome diversity between CDT positive and negative patients.

**Conclusions:** Patients with fecal samples that were positive for CDT had increased disease severity and worse clinical outcomes. Utilization of PCR and *C. difficile* Toxins A and B testing may not reveal the entire picture when diagnosing CDI and the detection of CDT-expressing strains may be valuable during patient treatment.

**Summary:** We found that CDI patients with CDT detected in stool by immunoassay were more likely to have severe disease and had higher 90-day mortality.

## Introduction

*Clostridioides difficile* (*C. difficile*) is a gram-positive, spore-forming bacteria that can cause diarrhea and colitis in patients with dysbiosis. In 2017, *C. difficile* infection (CDI) affected over 200,000 people a year, resulting in almost 13,000 deaths and was considered an urgent antibiotic resistant threat by the Centers for Disease Control (1). The increased prevalence of CDI is attributed to novel hypervirulent strains that cause worse disease and higher mortality (2). In addition to the main virulence factors, toxins A and B, these hypervirulent strains produce a third toxin known as *C. difficile* binary toxin (CDT) or binary toxin. CDT is comprised of CdtA, an actin-specific ADP-ribosyl transferase, and CdtB, the receptor-binding component (3). Our lab has shown that in a mouse model, CDT induces Toll-like receptor 2 (TLR2) pathways resulting in increased host inflammation and suppressed protective eosinophils (4).

With the increased prevalence of CDI in the United States, there has been substantial debate over the optimal diagnostic approach (5). The use of highly sensitive polymerase chain reaction (PCR) (that detects the presence of *C. difficile* but not toxin production) and increased clinical testing of asymptomatic individuals is believed to contribute to the overdiagnosis of CDI (6-8). While PCR positive, toxin A/B enzyme immunoassay (EIA) positive patients have increased disease severity compared to patients with discordant tests (PCR-, toxin A/B EIA+), fatal infections may occur even in the absence of detectable toxins A/B (8-10). It is uncommon for *C. difficile* strains to be ribotyped or tested for CDT-encoded genes, leading to insufficient information on potential prognosis. Previous studies have shown that detection of CDT genes in stool by PCR has been associated with worse outcomes, but no investigation has been done to compare disease severity and toxin detection by immunoassay (2, 11, 12).

Although little has been published on expressed CDT in stool, Carman et al (2018) showed that 027, by far the most dominant of the CDT-producing ribotypes, was about 10-fold more abundant in patient feces than was 014/020 and made about 10-fold more TcdA and TcdB than 014/020. Ribotype 014/020 is a non-CDT producer and probably the most widespread and abundant ribotype before the emergence of fluoroquinolone resistance, notably in 027. Furthermore 027 infection resulted in about a five-fold increase in fecal lactoferrin compared with 014/020 (13). In this study, we wished to further explore the relationship between CDT and disease severity by testing for toxin in stool with enzyme-linked immunosorbent assay (ELISA). We found that CDI patients with CDT-expressing strains had increase disease severity and worse clinical outcomes.

## Methods

### Sample Collection and Patient Information

Stool collected from 215 CDI patients were retrospectively identified from the University of Virginia (UVA) Medical Center’s electronic medical records (EMR). Stool samples were collected between 2015-2017 and were held in the clinical laboratory at 4°C for 48 hours, then aliquoted and stored at -80°C until testing. All patients had diarrhea and were positive for *C. difficile* by Xpert *C. difficile* PCR (Cepheid, Sunnyvale, CA). Data on the presence or absence of CDT genes was not collected, nor were isolates collected for typing. Patient demographics, clinical lab results and disease outcomes were collected retrospectively from UVA Clinical Data Repository and the EMR. The collection of patient data was approved by the institutional review board (protocol IRB-HSR 16926).

### CdtB Detection

Stool samples from 215 patients were tested using a previously described research-only ELISA for Clostridium difficile CdtB (14).

### *C. difficile* TcdA and TcdB Detection

All stool samples were tested for **TcdA and TcdB** by C. DIFFICILE TOX A/B II™ according to manufacturer instructions (catalog no. T5015, TechLab Inc, Blacksburg, VA).

### Lactoferrin Detection

Stool lactoferrin was assayed using LACTOFERRIN SCAN® according to manufacturer instructions (catalog no. T5009 / 30351, TechLab Inc, Blacksburg, VA).

### Fecal DNA Extraction/16S rRNA Gene V4 region sequencing

DNA from patient fecal samples was extracted using the QIAamp DNA Stool Mini Kit (catalog no. 51504, Qiagen, Hilden, Germany). For each sample, the V4 region of the 16S rRNA gene was amplified using the dual indexing sequencing strategy as described previously (15). Sequencing was done on the Illumina MiSeq platform, using a MiSeq Reagent Kit V2 500 cycles (Illumina cat# MS102-2003), according to the manufacturer’s instructions with modifications found in the Schloss SOP: https://github.com/SchlossLab/MiSeq_WetLab_SOP. The mock community ZymoBIOMICS Microbial Community DNA Standard (Zymo Research cat# D6306) was sequenced to monitor sequencing error.

### 16 S rRNA Gene Amplicon Curation and Analysis

The 16S data curation and analysis from the human stool samples was performed using R version 4.0.3. Sequences were curated using the R package DADA2 version 1.18 (16). Briefly, reads were filtered and trimmed using standard parameters outlined in the DADA2v 1.8 pipeline. The error rates for the amplicon datasets were determined using the DADA2’s implementation of a parametric error model. Samples were then dereplicated and sequence and variants were inferred. Overlapping forward and reverse reads were merged and sequences that were shorter than 251 bp or longer than 255 bp were removed. Finally, chimeras were removed. Taxonomy was assigned to amplicon sequence variants (ASVs) using the DADA2-formatted SILVA taxonomic training data release 132 (17). Samples with less than 16,000 reads per sample were removed from the analysis (this included the extraction blank). The sequences associated with this analysis will be deposited in the SRA. Following sequence curation, the packages phyloseq v1.34.0, vegan v2.5.7, dplyer v1.0.6 and ggpubr v0.4.0 were used for analysis and generation of figures (18).

### Statistical Methods

All statistical comparisons and graphs were made using GraphPad Prism 9 or using R version 4.0.3. Comparisons of CDT positive and CDT negative patient demographics, Toxin A/B status and disease outcomes were calculated using Chi-square test. Differences in white blood cell counts (WBC), clinical PCR cycle threshold (Ct) values, stool lactoferrin, days in the ICU and days hospitalized between CDT positive and CDT negative patients were calculated using a Mann-Whitney U test. Differences between Simpson and Shannon indexes were found using a Wilcoxon signed-rank test. A p-value greater than 0.05 was considered significant.

## Results

### Patient Characteristics

Of the 215 patient stool samples analyzed, 32 were positive for CDT protein (Table 1). Nine of the 32 were negative in the A/BII ELISA. Both CDT positive and CDT negative cohorts were split evenly by sex, and there was no significant difference in age (p=0.1005). CDT positive patients were more likely to also be positive for Toxins A/B (p = 0.0006) and have a WBC count higher than 15,000/μl (p=0.0019). Patients testing positive for CDT were also more likely to be admitted to the ICU (p=0.0113) and more likely to die within 90 days of diagnosis (p=0.0320).

**Table 1.** Patient Characteristics.

| Characteristic | CDT+ (32) | CDT- (183) | p value |
| --- | --- | --- | --- |
| **Age** (mean, SD) | 64.7 (16.2) | 59.0 (17.0) | 0.1005 |
| **Sex** (No, %)* |  |  | 0.9258 |
| Female | 16 (50) | 85 (50.1) |  |
| Male | 16 (50) | 82 (49.9) |  |
| **Toxin A/B+** (No, %) | 23 (71.9) | 72 (39.3) | **0.0006** |
| **WBC > 15,000/μl** (No, %) * | 14 (45.2) | 34 (19.5) | **0.0019** |
| **Admitted to ICU** (No, %) * | 11 (34.4) | 28 (15.6) | **0.0113** |
| **Deceased within 90 days of diagnosis** (No, %) * | 7 (23.3) | 16 (9.6) | **0.0320** |

\*Some clinical data missing, percentage based on patients with data available

### Patients testing positive for CDT had increased bacterial burden, white blood cell count and intestinal inflammation

The 32 CDT positive patients had significantly higher WBC (p=0.0129, Figure 1a). These patients also had higher bacterial burden, as indicated by lower Ct value (p=0.0008, Figure 1b). Of the 32 CDT positive patients, 9 were toxin A/B negative. When we compared bacterial burden from the samples that were discordant for the three toxins (CDT positive, negative for toxin A/B) to the burden in the 23 patients that tested positive for both CDT and toxins A/B, those that were positive for all toxins had significantly lower Ct values (Figure S1, p=0.0096). Thus, patients with CDT positive, toxin A/B positive samples had higher *C. difficile* burden compared to patients positive for CDT and negative for toxin A/B. Patients testing positive for CDT trended to have higher lactoferrin levels compared to CDT negative patients (p=0.0646, Figure 1c).

**Figure 1.**
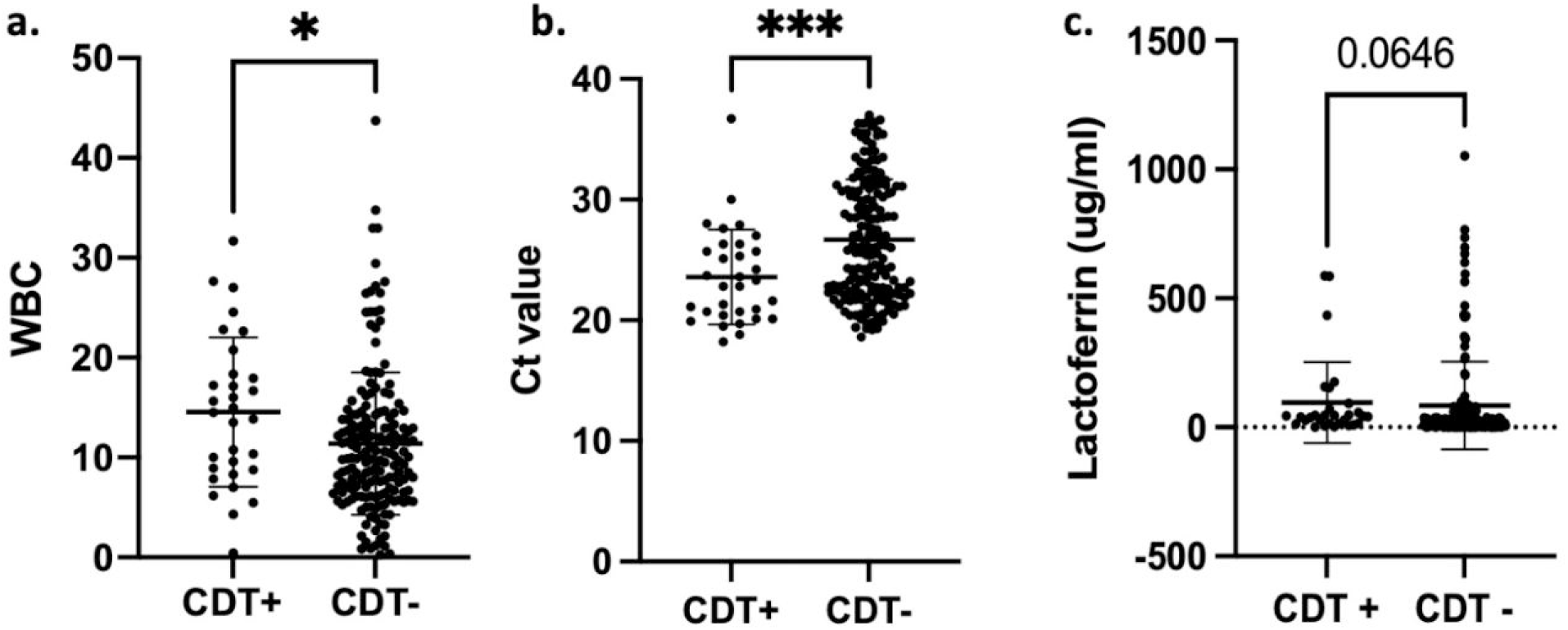
Patients testing positive for CDT had increased bacterial burden, white blood cell count and intestinal inflammation. (a) Patients with CDT-expressing strains had higher white blood cell counts than patients with non-CDT expressing strains (p=0.0129). (b) CDT positive patients had lower Ct values than CDT negative patients, indicating increased bacterial burden (p=0.0008). (c) CDT positive patients had elevated lactoferrin comparted to CDT negative patients, although not statistically significant (p=0.0646)

### CDT positive patients had increased hospital and ICU stays

Of the 215 CDI patients, the CDT positive patients had longer hospitalizations (p=0.0468) and days in the ICU (p=0.0083, Figure 2). Patients testing positive for CDT stayed in the hospital 6.5 days longer than CDT negative patients. CDT negative patients stayed in the ICU on average 1.73 days compared to 3.59 days for CDT positive patients.

**Figure 2.**
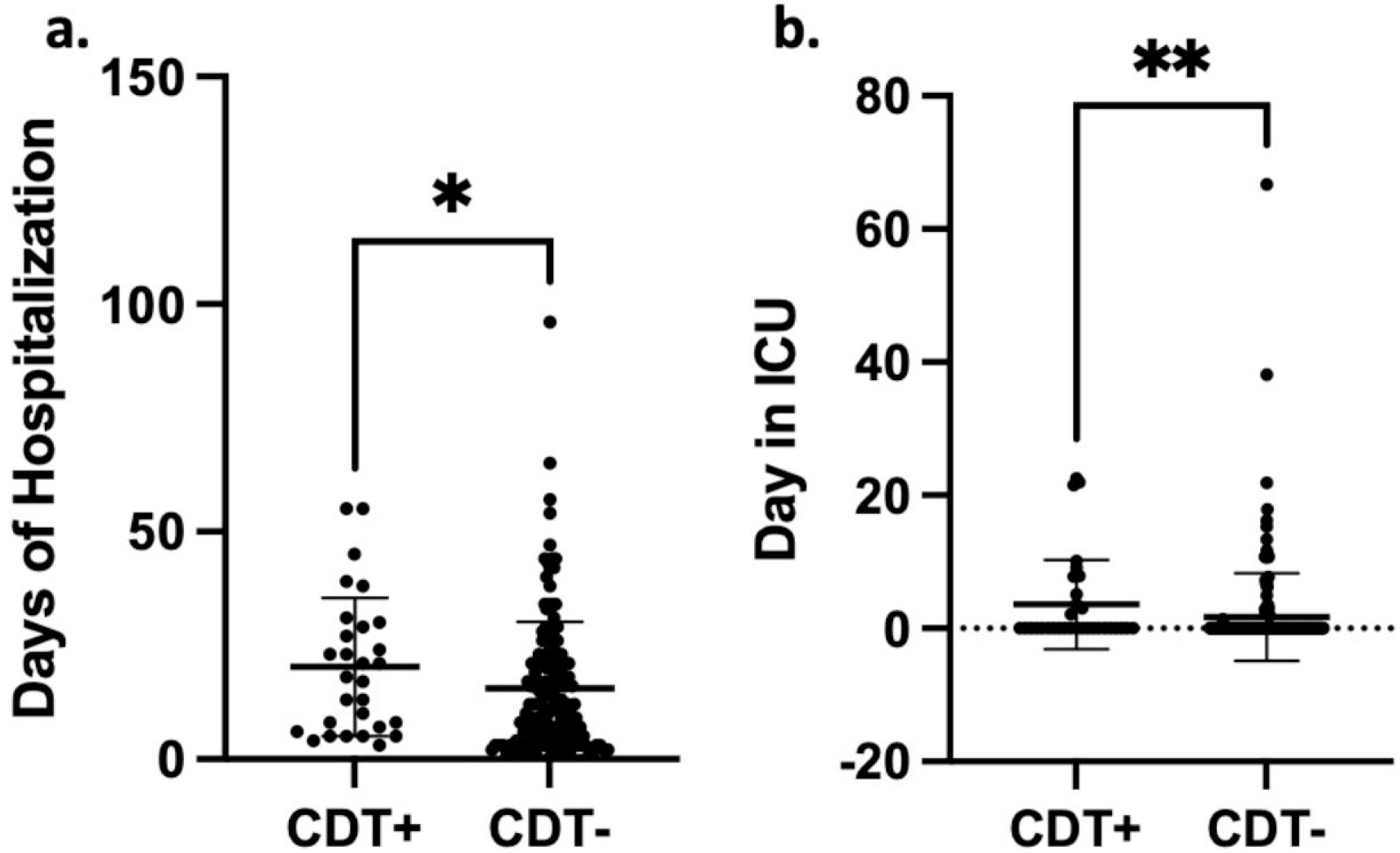
CDT positive patients had increased hospital and ICU stays. (a) Patients with CDT detected in their stool had longer stays in the hospital, with an average stay of 18 days compared to 11.5 days for CDT negative patients. (p=0.0468). (b) CDT positive patients had longer stays in the ICU (p=0.0083).

### No differences in gut microbiome alpha diversity between CDT positive and negative patients

We sought to determine if the structure of the fecal microbiome was altered in CDT+ patients. Composition of the fecal microbiota from 172 patients was profiled using amplicon sequencing of the 16S rRNA gene V4 region. While CDT negative patients trended towards slightly increased alpha diversity as measured by either Simpson diversity index or Shannon diversity, there was not a significant difference using either metric (Figure 3a, p=0.26, Figure 3b, p=0.16).

**Figure 3.**
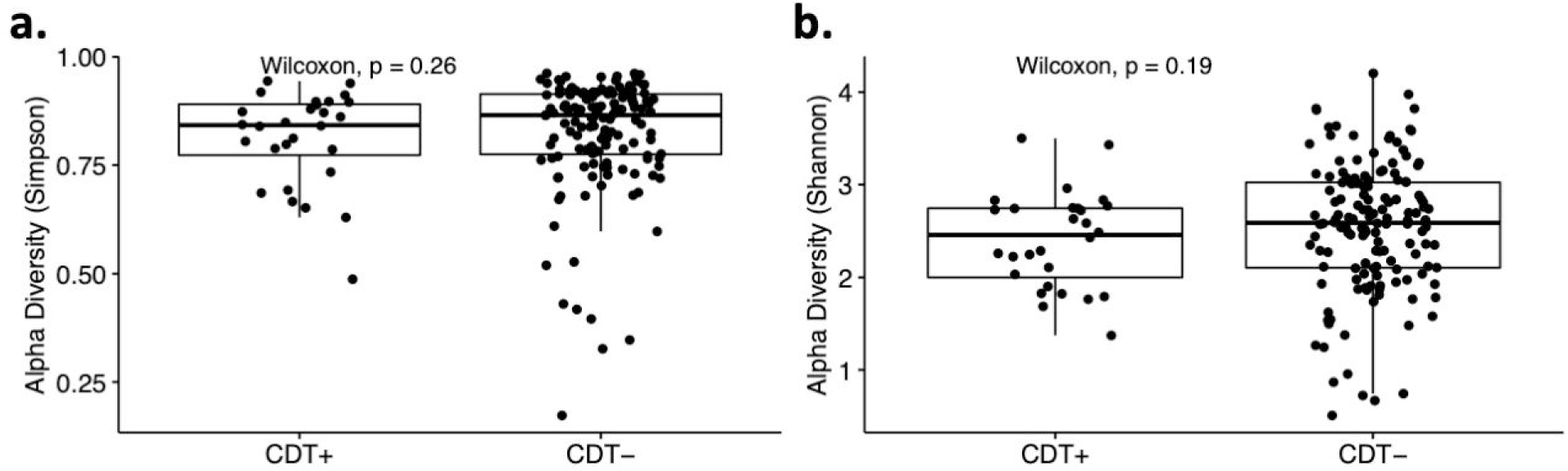
No differences in gut microbiome diversity between CDT positive and negative patients. (a) There was no significant difference in Simpson Diversity Index between CDT positive and negative patients (p=0.26). (b) Shannon Diversity Index was slightly higher in CDT negative patients although not statistically significant (p=0.16).

## Discussion

Previous studies have shown that infection with strains positive for the CDTb gene result in increased disease severity, increased disease recurrence and higher mortality rates (2, 11-13). Here we reiterate those earlier findings and further show that patients with CDT protein present in stool (9/32 (28%) of which had no detectable toxin A/B production) had worse disease outcomes, with increased rates of ICU admissions and higher mortality. Additionally, these patients had longer stays in the hospital and ICU, causing increased burden to health care systems.

Other measures of disease severity were also worse in CDT+ infections. One characteristic of severe CDI is having a WBC count of greater than 15,000/μl (19). We found that 45.2% of CDT positive patients were considered to have severe CDI based on this guideline compared to 19.5% of CDT negative patients. Overall CDT positive patients had higher WBC counts (p=0.0129). Other groups have similarly found that patients infected with *C. difficile* strains encode for CDT have elevated WBC count (2, 11, 12). We did not find any significant differences in neutrophils, eosinophils, lymphocytes, monocytes, or basophils, although our data set was incomplete for these values. Patients with CDT positive stools also exhibited higher bacterial burdens as indicated by lower Ct values (p=0.0008) consistent with higher burden being correlated with more severe disease (13, 20). We also found that CDT positive, toxin A/B positive patients had lower Ct values compared to CDT positive, toxin A/B negative (p=0.0096), even though measures in clinical disease did not differ. Boone et al found that patients with the CDT producing hypervirulent ribotype 027 had increased lactoferrin compared to patients colonized with other strains (21). We confirmed the earlier finding that CDT positive patients had higher lactoferrin levels, although in our instance this was not statistically significant (p=0.06).

Lastly, we performed 16 S rRNA gene sequencing to determine if there were differences in gut microbiome diversity in CDT positive and negative patients. Studies have described a decreased gut microbiome diversity in patients with CDI compared to control patients (22-24). We found no significant differences in Simpson or Shannon indexes, although CDT positive patients tended towards having lower microbiome diversity. These results indicate that increased disease severity related to CDT is not associated with decreased gut microbiome diversity. In conclusion, we believe we are the first to demonstrate that patients with stool testing positive for CDT by ELISA have more severe CDI and worse disease outcomes. The presence of stool CDT was associated with higher bacterial burden and WBC counts, as well as longer hospital and ICU stays and increased mortality. There was no significant difference in gut microbiome diversity between CDT positive and negative patients. Testing for the presence of CDT in stool is a valuable tool to better explain differences in disease severity amongst CDI patients. Detection of CDT and CDT-expressing strains may be beneficial in disease diagnosis and treatment.

## Data Availability

We will submit all corresponding data and code to Dryad or NPG hub upon the acceptance of our submission.

## Acknowledgments

We would like to thank Jennifer White for IRB protocol preparation, and Research & Clinical Trial Analytics, Ken Scully, and Li Mo for clinical data retrieval.

---

**Supplemental Figure 1. Higher bacterial burden in CDT positive, toxin A/B positive patients compared to patients positive for CDT and negative for toxin A/B**. CDT and toxin A/B positive (n=23) had lower Ct values compared to CDT positive, toxin A/B negative patients (p=0.0096)

