## Supplementary figures and images for "Binary toxin expression by *Clostridioides difficile* is associated with worse disease"

### Supplemental Figure 1

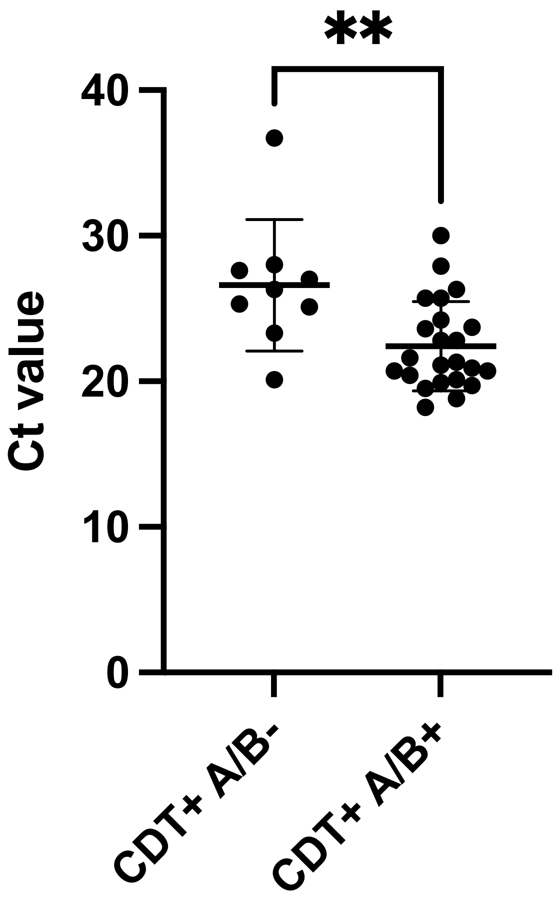
